# Hydroxychloroquine/Chloroquine in COVID-19 With Focus on Hospitalized Patients – A Systematic Review

**DOI:** 10.1101/2022.01.11.22269069

**Authors:** Daniel Freilich, Jennifer Victory, Anne Gadomski

**Affiliations:** Bassett Medical Center; Bassett Research Institute

**Keywords:** Hydroxychloroquine, chloroquine, COVID-19, SARS-CoV-2, clinical trials, in vitro, animal studies

## Abstract

**Background:** In the beginning of the COVID-19 pandemic, many hospitalized patients received empiric hydroxychloroquine/chloroquine (HC/CQ). Although some retrospective-observational trials suggested potential benefit, all subsequent randomized clinical trials (RCTs) failed to show benefit and use generally ceased. Herein, we summarize key studies that clinicians advising patients on HC/CQ’s efficacy:safety calculus in hospitalized COVID-19 patients would want to know about in a practical one-stop-shopping source.

**Methods:** Pubmed and Google were searched on November 4, 2021. Search words included: COVID-19, hydroxychloroquine, chloroquine, *in vitro*, animal studies, clinical trials, and meta-analyses. Studies were assessed for import and included if considered impactful for benefit:risk assessment.

**Results:** These searches led to inclusion of 12 *in vitro* and animal reports; 12 retrospective-observational trials, 19 interventional clinical trials (17 RCTs, 1 single-arm, 1 controlled but unblinded), and 51 meta-analyses in hospitalized patients.

Inconsistent efficacy was seen *in vitro* and in animal studies for coronaviruses and nil in SARS-CoV-2 animal models specifically. Most retrospective-observational studies in hospitalized COVID-19 patients found no efficacy; QT prolongation and increased adverse events and mortality were reported in some. All RCTs and almost all meta-analyses provided robust data showing no benefit in overall populations and subgroups, yet concerning safety issues in many.

**Conclusions:** HC/CQ have inconsistent anti-coronavirus efficacy *in vitro* and in animal models, and no convincing efficacy yet substantial safety issues in the overwhelming majority of retrospective-observational trials, RCTs, and meta-analyses in hospitalized COVID-19 patients. HC/CQ should not be prescribed for hospitalized COVID-19 patients outside of clinical trials.

**Key Summary Points:** Preclinical hydroxychloroquine/chloroquine *in vitro* studies found inconsistent activity against coronaviruses including SARS-CoV-2.

Preclinical hydroxychloroquine/chloroquine animals studies found inconsistent efficacy for coronaviruses in general and none for SARS-CoV-2.

The overhwelming majority of RCTs and retrospective-observational trials found no benefit for hydroxychloroquine/chloroquine in hospitalized COVID-19 patients, and many found concerning safety signals.

The majority of RCTs and retrospective-observational trials found no benefit for hydroxychloroquine/chloroquine in COVID-19 outpatients or for pre- or post-exposure prophylaxis, and some found concerning safety signals.

The overwhelming majority of meta-analyses found no benefit for hydroxychloroquine/chloroquine in COVID-19 inpatients, outpatients, or for prophylaxis, and many found concerning safety signals.

## Introduction

More than 770,000 Americans have died of COVID-19, and U.S. deaths continue at 1,000-2,000 daily. Thus, it is imperative that we confirm the beneficial efficacy:safety calculus of effective medications (positive studies) but also the absence of beneficial efficacy:safety calculus of purportedly effective medications (negative studies), so we refrain from prescribing them potentially causing harm. Lessons should be learned from the successes and failures in response to the pandemic. The story of the medical community’s empiric prescribing of hydroxychloroquine (HC) and chloroquine (CQ) despite weak *a priori* data and a progressively negative preclinical and clinical database during the pandemic should be instructive to avoid repetition in this latter phase of the pandemic.

HC received early pandemic attention for use in COVID-19 in part because then President Trump recommended and took it for post-exposure prophylaxis. HC was frequently administered empirically (peaking at a whopping 42% prevalence for hospitalized COVID-19 patients early in the pandemic [1]) and recommended in some expert reviews and guidelines (e.g., [2]). Supportive data were shaky, relying on inconsistent *in vitro* and animal studies mainly for other coronaviruses [3-4], and small flawed uncontrolled trials not confirming benefit [5-6]). Notwithstanding, the FDA issued an Emergency Use Authorization (EUA) in March 2020 for HC/CQ use in hospitalized COVID-19 patients who could not be enrolled in clinical trials [7].

Retrospective observational studies were subsequently published with the overwhelming majority finding no benefit and some finding higher mortality and toxicities (e.g., QTc prolongation) [6, 8-25], but they were fraught with potential bias, in part because HC/CQ patients were usually sicker. Most American guidelines recommended use in COVID-19 only in trials.

In late spring/early summer, 2020, results of at least 5 RCTs became available [26-30], with all showing no primary outcome benefit in hospitalized COVID-19 patients (and safety concerns in most). Most trialists studying treatments for hospitalized COVID-19 patients discontinued enrollment in their HC/CQ study arms by the summer of 2020. Twelve additional RCTs later in 2020, and in 2021, found similar results [31-42]. Numerous meta-analyses were completed assessing HC/CQ in hospitalized COVID-19 patients – almost all found ineffectiveness, and many found adverse safety signals (e.g., [43-45]).

This review summarizes the evolved benefits:risks database for HC/CQ in the COVID-19 pandemic, aiming to assist clinicians in easily summarizing the evidence basis for their benefits:risk assessment to their patients, some of whom (as well as other experts) continue to believe in possible effectiveness of these medications in some indications (e.g., [46]).

## Methods

Pubmed and Google searches were conducted, with the latest repeated November 4, 2021. Initial search words included “COVID-19”, “hydroxychloroquine”, and “chloroquine”, found 3,123 publications.

Adding search words, “clinical trials”, reduced this list to 78 publications – 15 RCTs and 5 retrospective-observational trials in hospitalized patients, 4 RCTs in outpatients, 6 RCTs evaluating prophylaxis, and 48 other studies (not evaluating HC/CQ, not trials, unable to categorize). For hospitalized COVID-19 patients, the primary focus of this review, the Google search, as well as findings from the other Pubmed searches below (e.g., *in vitro* studies), led to inclusion of 4 additional studies (2 RCTs, 1 single-arm trial, and 1 prospective controlled observational trial) and 7 additional retrospective-observational trials yielding 19 interventional trials (17 of which were RCTs) and 12 retrospective-observational trials. Of the 17 RCTs, 5 published earlier in the pandemic were evaluated in detail systematically because they impacted evolving FDA recommendations and guidelines more than latter RCTs which tended to be confirmatory and additive to established literature (summarized but not detailed herein); all 5 included hard primary outcomes (i.e., mortality, ordinal scores, and viral clearance) with most comparing primary outcome risk, rate, and odds ratios; standard baseline patient characteristics were collected in all; 4 vs. 1 were assessed as having moderate and mild risk of bias, respectively (Table S1).

Adding “meta-analysis” to the Pubmed search led to 62 publications, 12 of whom were not focusing on hospitalized COVID-19 patients were excluded yielding a total of 50 meta-analyses; our unpublished IPD meta-analysis was added yielding a total of 51 meta-analyses in hospitalized patients (3 examples were detailed).

The Pubmed search led to the finding of 4 outpatient RCTs; the Google search found two additional studies (1 retrospective-observational study and 1 RCT) yielding a total of 6 outpatient studies.

The Pubmed search led to finding 6 prophylaxis studies; the Google search found an additional 3 studies (1 RCT, 2 observational cohort) yielding a total of 9 prophylaxis studies.

Adding the key words, “in vitro”, to our Pubmed search, resulted in 277 publications of which only 6 were in fact *in vitro* studies and thus included in our preclinical review. Our Google and other Pubmed searches found 2 additional studies, yielding 8 *in vitro* studies. Adding key words, “animal studies”, resulted in 89 publications of which only 4 were in fact animal studies and thus included in our preclinical summary; no additional studies were found with the Google or other Pubmed searches. In total, 12 preclinical studies were included in this review (8 *in vitro*, 4 animal).

See Figure 1.

**Figure 1.**
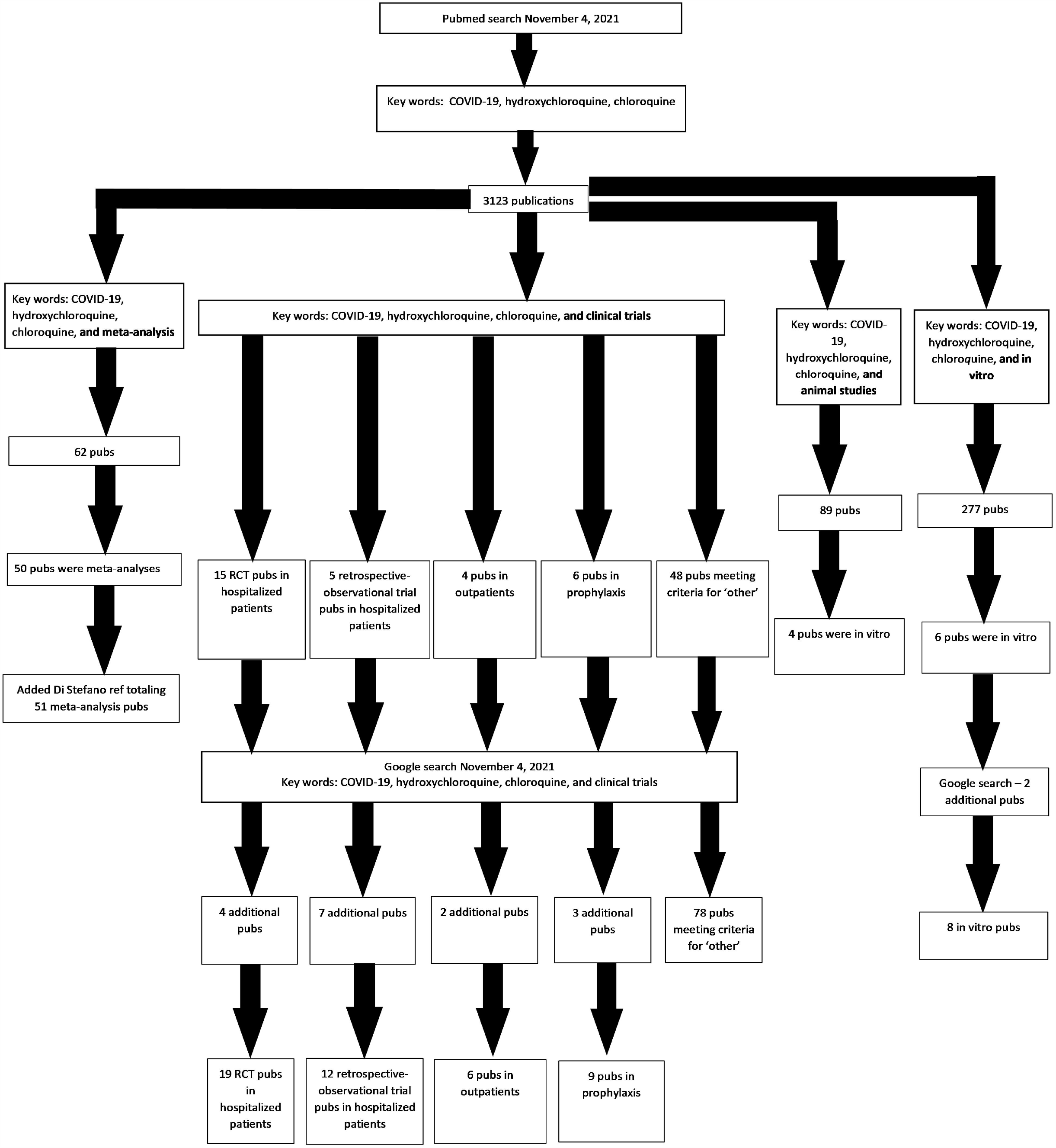
Pubmed and Google searches were used to query the published preclinical and clinical literature for hydroxychloroquine and chloroquine use for treatment and prophylaxis in COVID-19

We prioritized detailing of RCTs, larger retrospective-observational studies, and larger (especially IPD) meta-analyses as these were considered ‘more impactful’. Uncontrolled trials, smaller retrospective-observational studies, and smaller aggregate data meta-analyses (and as noted, later confirmatory RCTs) were considered ‘less impactful’.

This systematic review was not based on a written protocol nor was it registered.

## Results and Discussion

### Preclinical *In vitro* Studies

The anti-inflammatory and antimalarial medications, HC and CQ, demonstrated antiviral activity against SARS-CoV and MERS Co-V *in vitro*, including primate cells – although inconsistently as no activity was seen in SARS-CoV mouse cell culture [47].

A handful of reports showed SARS-CoV-2 growth inhibition *in vitro* by HC/CQ [48-50] including synergy with azithromycin at clinically realistic concentrations [51] – albeit similarly inconsistently as only two of three studies found viral growth inhibition in Vero E6 cell (African green monkey kidney cells) and one found no growth inhibition in a human airway epithelial model [52-54]. One study found that growth inhibition by quinine exceeds that by HC or CQ in Vero cells, human Caco-2 colon epithelial cells, lung A549 cells, and Calu-3 lung epithelial cells [55].

The mechanism of action of HC/CQ’s antiviral activity is purported to be via enhancement of endosomal pH leading to decreased viral-cell fusion and inhibition of glycosylation of SARS-CoV cellular receptors (both leading to decreased cell entry) [56]; immune modulation and anti-thrombotic characteristics may also be important. CQ EC50s of 0.77-6.9 microM have been reported, levels reached in patients receiving HC for rheumatoid arthritis [48, 12, 57]. One study reported an EC50 5.47 microM for CQ vs. 0.72 for HC for SARS-CoV-2 [58].

Overall, HC/CQ have *in vitro* activity against coronaviruses including SARS-CoV-2 albeit inconsistently and not as potently as some other antivirals [59].

### Preclinical *In Vivo* Animal Studies

Animal studies evaluating HC/CQ for coronaviruses have been inconsistent. In mice, HC activity was found against the human coronavirus HCoV-OC43 [60] but none against SARS-CoV [47].

The limited database of animal studies with SARS-CoV-2 has been relatively ‘negative.’ Neither standard nor high dosing of prophylactic or therapeutic HC was efficacious in hamsters; standard prophylactic and therapeutic dosing was similarly ineffective in rhesus macaques [3]. Clinical parameters, viral shedding/load, and lung pathologic changes were similar in treatment and control groups. Another hamster study also showed no treatment or prophylaxis efficacy for HC [61]. In a ferret SARS-CoV-2 model, HC marginally decreased clinical scores at some time points but had no effect on symptoms duration or viral shedding or load [4]. In a macaque SARS-CoV-2 model, treatment dosing with HC with or without azithromycin had no effect on viral clearance, and prophylactic dosing did not decrease infection [54].

Notably, in other viral infections (e.g., influenza), CQ failed to replicate promising *in vitro* findings in *in vivo* animal and human studies [62].

The *in vitro* data for SARS viruses and relative safety of these medications in malaria and rheumatoid arthritis in part prompted recommendations by some expert groups and guidelines early in the pandemic to administer these drugs empirically (ideally in RCTs) in COVID-19 [2, 63).

### Retrospective-Observational Clinical Studies in Hospitalized Patients

Early in the pandemic, a small open-label non-randomized French trial was published, reporting results for HC treatment of hospitalized COVID-19 patients [8]. This pilot study received undue attention beyond its scientific and clinical significance, being referenced by then President Trump. Patients were given HC with or without azithromycin; patients from other hospitals and those refusing participation served as negative controls. The primary outcome measurement, day-6 virologic clearance by nasopharyngeal swab PCR, occurred in 70% of HC vs. 12.5% of control patients (p=0.001) (100% in HC/azithromycin patients and 57.1% in HC patients [p<0.001]); Six of 42 enrolled patients were lost to follow-up in the HC group. Mean serum HC concentration was 0.46 mcg/ml±0.2, akin to EC50s published for CQ for SARS-CoV viruses. The trial was limited by small size, open-label design without true controls, short follow-up, and high dropout. The authors concluded their results were “*promising*” and recommended HC with azithromycin in COVID-19. In a follow-up reanalysis, the authors found that their results for HC/azithromyin were similar even after addressing critiques about excluded patients and outcome adjudication [8]. These results, however, could not be replicated in a subsequent study by other investigators [6].

Another small Chinese study early in the pandemic found improved clinical outcomes with HC in hospitalized COVID-19 patients [9]. The study randomized 62 patients to standard care with or without HC for 5 days in a double-blinded fashion. Time to Clinical Recovery, the primary outcome, was significantly shorter with HC. Improvement in pneumonia by CT imaging was higher with HC (80.6% vs. 54.8%).

In another early pandemic uncontrolled retrospective-observational study focused on hospitalized mechanically ventilated COVID-19 patients, also from China, mortality was 9/48 (18.8%) with HC vs. 238/502 (47.4%) without HC (p<0.0001) [10].

These small studies suggested that HC might have efficacy in COVID-19. Additional Chinese RCTs early in the pandemic led to the inclusion of recommendation in some Chinese guidelines to treat hospitalized COVID-19 patients with HC/CQ [11] after data from more than 100 patient also showed less pulmonary complication and more rapid viral shedding and clinical improvement [12].

Based mainly on the limited preclinical data and these small early RCTs, in part, the FDA issued an EUA for HC/CQ for treatment of hospitalized COVID-19 patients for whom enrollment in trials was impractical on March 28, 2020 [7].

A small Brazilian trial was then published [13-14] demonstrating higher QTc prolongation rates (18.9% vs. 11.1%) and higher mortality (39% vs. 15% [OR, 3.6; 95% CI, 1.2-10.6]) with higher vs. lower CQ dosing for 81 hospitalized COVID-19 patients. There was no difference in viral shedding clearance. Prolonged QTc was not associated with death and no torsades de pointes occurred. Limitations included absence of placebo control and published mitigation strategies to reduce QTc prolongation (e.g., excluding baseline QTc prolongation and co-administration of additional QTc prolonging medications [100% received azithromycin]), single-center design, small sample size, and baseline imbalance.

Prolonged QTc was also reported in two additional retrospective case series of hospitalized COVID-19 patients treated with HC with or without azithromycin. In the small French series of 40 patients, baseline QTc>460 msec was an exclusion. 93% developed some QTc prolongation; 36% developed more severe QTc prolongation (more commonly with concomitant azithromycin [33% vs. 5%, p=0.03]). No ventricular arrhythmias including torsades de pointes occurred. Seven patients (17.5%) stopped medication due to adverse events, ECG changes, or acute renal failure [15]. In the Boston series in 90 patients, combined therapy was associated with a larger median increase in the QT interval than HC monotherapy (23 vs. 5.5 msec, p=0.03), resulting in 13% vs. 3% of patients, respectively, having QTc change >/= 60 msec. The risk of QTc prolongation to >/=500 msec was similar (21% vs. 19%). The authors implied a baseline QTc prolongation exclusion. Ten patients (11%) discontinued medication because of adverse events (nausea, hypoglycemia, and one case of delayed torsades de pointes) [16].

None of these series had control arms, so the relative risk of QTc prolongation remained elusive. Yet, it appeared that higher dosing and co-administration of QTc prolonging medications resulted in more frequent and severe QTc prolongation. Torsades de pointes, the feared QTc prolongation complication, appeared rare (occurring in only 1 of 211 patients in the series [0.5%]). An accompanying *JAMA* editorial concluded, the studies “*underscore the potential risk … of hydroxychloroquine* … *It is also true that … the QTc can be safely monitored in most patients*” [64].

Based in part on these safety issues, the FDA issued a *Drug Safety Communication* on 24 April 2020 reminding providers about HC/CQ risks in COVID-19, mitigation strategies, and warning against use in outpatients and outside trials [65]. NIH COVID-19 guidelines in April 2020 concluded there remained equipoise for these drugs, and risk mitigation strategies should be employed if used. Some experts disagreed with the FDA’s allowance for continued empiric use despite emerging efficacy lapse yet concerning safety issues.

A retrospective study on 368 Wisconsin VA hospitalized COVID-19 patients compared mortality and mechanical ventilation with HC with or without azithromycin or neither. The study found higher mortality with HC vs. no HC (27.8% vs. 11.4%: adjusted HR, 2.61; 95% CI, 1.1-6.17, p=0.03) but no mechanical ventilation difference [17]. The study was small, retrospective/observational, and without randomization; HC patients were sicker.

Effects on mortality and intubation were equivocal in two large New York retrospective observational studies published in May 2020 [18-19] The Columbia University study compared a composite outcome of intubation and death in 1,376 hospitalized COVID-19 patients treated with HC or not and found no significant associations in crude, multivariable, and propensity-score analyses [18]. The second study was in 1,438 hospitalized COVID-19 patients from 25 hospitals in NY State treated with HC with or without azithromycin or azithromycin alone [19]. In-hospital mortality was not statistically different, 25.7%, and 19.9% for HC with and without azithromycin, respectively, and 10% for azithromycin. More frequent cardiac arrest was found in patients receiving both drugs. No ECG abnormalities differences were found. HC patients were sicker in both studies.

After the NY studies, *Lancet* published the largest retrospective observational study to date, comparing in-hospital mortality and arrhythmias in 96,032 hospitalized COVID-19 patients treated with HC (or CQ) with or without azithromycin vs. neither [20]. In-hospital mortality and arrhythmias occurred significantly more frequently with HC/CQ, especially with azithromycin, in all analyses. Although larger, this study was limited by the same observational/retrospective confounding as the prior studies. Although the authors reported similar between-group baseline characteristics, others found HC/CQ patients to be sicker. The publication was retracted. WHO temporarily halted HC arm enrollment in its *Solidarity* Trial after this publication.

Another French study in hospitalized COVID-19 patients (focusing on those requiring oxygen) compared mortality in 84 HC vs. 89 control patients. 21-day survival without ICU transfer was 76 vs. 75%, respectively (weighted hazard ratio 0.9, 95% CI 0.4 to 2.1). Eight HC patients (10%) developed arrythmias of which 7 were QTc prolongation (vs. 0 in control patients) [21].

In May 2020, the FDA published *Pharmacovigilance Memorandum Safety* data from its *Adverse Event Reporting System* (*FAERS*) and other sources [66]. QT prolongation was the most common cardiac SAE, with co-administration of other QT-prolonging medications occurring in most cases; other cardiac SAEs included torsades de pointes in 4%, ventricular arrhythmia in 13%, and death in 23%. The most common non-cardiac SAE was increased LFTs. Four unexpected methemoglobinemia SAEs occurred.

A Weill Cornell Medicine (New York City) single-arm HC study in 153 patients found improvement in hypoxia scores in 52%, no ventricular arrhythmias, and QTc prolongation leading to drug discontinuation in 2% [23].

A Henry Ford Health System (southeast Michigan) study was reported ^[22]^, comparing in-hospital mortality in 2,541 hospitalized COVID-19 patients treated with HC (13.5% [95% CI: 11.6-15.5%]), HC with azithromycin (20.1% [95% CI: 17.3%-23.0%], azithromycin 22.4% [95% CI: 16.0%-30.1%], and standard care 26.4% [95% CI: 22.2%-31.0%]. HC with or without azithromycin led to hazard ratio mortality reduction of 66-71%. Of all the larger retrospective trials, this study stands out as a positive one; however, it too was observational, without randomization or blinding, and confounded (e.g., steroids were given to 74.3-78.9% of HC vs. 35.7-38.8% of non-HC patients).

Two additional relatively small retrospective-observational studies reported decreased mortality with HC monotherapy and with co-administration with azithromycin [24-25]. In a single-site retrospective cohort study hospitalized patients with COVID-19 pneumonia, deaths occurred in 102/297 HC + azithromycin patients (34.3%) vs. 7/17 HC alone patients (41.2%) vs. 35/63 patients receiving neither due to ‘contraindications’ (55.6%). Use of HCQ + azithromycin (vs. no treatment) was inversely associated with inpatient mortality HR 0.265 (95% CI 0.171-0.412, P<0.001) [24]. A preprint of an observational study from early in the pandemic in 255 hospitalized mechanically ventilated patients at a single New Jersey site reported a logistics regression survival odds ratio of 14.18 (95% CI, 4.05-55.61, p<0.0001) in patients receiving HC and azithromycin [25].

These studies were observational, without randomization or blinding, and with baseline imbalances and other potential sources of confounding.

### RCTs in Hospitalized Patients

At last, results from at least five RCTs became available in the spring/summer of 2020 [26-30]. These are detailed below because they significantly impacted ensuing FDA and guidelines changes.

The first was a Chinese multicenter open-label RCT in 150 hospitalized laboratory-confirmed COVID-19 patients, 148 of whom had mild (negative chest x-ray) to moderate disease (positive chest x-ray) [26]. The mean interval from symptoms onset was 16.6 days. There was no significant difference in the main outcome measurement, intention-to-treat analysis of nasopharyngeal swab (SARS-CoV-2 PCR) negative conversion, which occurred in 85.4% of HC vs. 81.3% of standard care patients (difference 4.1%; 95% CI -10.3% to 18.5%). Adverse events (AEs) occurred in 30% vs. 9% (diarrhea most commonly), and SAEs occurred in 2 vs. 0 patients, respectively. No arrhythmias or QTc prolongation were reported. Study limitations included small size, delayed administration, early termination, open-label, other COVID-19 treatments, mild-moderate severity focus, and high dosing.

The second RCT was the *Recovery Trial* June 5, 2020, press release and eventual publication in *NEJM* on October 8, 2020 [27]. The pragmatic platform design included randomization but open-label, and standard care control without placebo – in 176 United Kingdom centers.^(36)^ The mean interval from symptoms onset was 9 days. The trial included 17% of patients with severe disease (requiring mechanical ventilation or ECMO), 60% with moderate disease (requiring oxygen or noninvasive ventilation), and 24% with mild disease (requiring neither). The study’s primary outcome measurement, 28-day mortality, was reached in 418/1,561 (26.8%) of HC vs. 788/3,155 (25%) of standard care patients (RR 1.09; 95% CI 0.96 to 1.23; p=0.18). Secondary outcomes included higher hospital length of stay (16 vs. 13 days, respectively), a higher composite endpoint of mechanical ventilation requirement and death (29.8% vs. 26.5%, respectively; RR 1.12; 95% CI 1.01-1.25), and higher stratified 28-day mortality trend in HC patients. Trial strengths included large size, randomization, control, and similar steroid use in both groups; limitations were open-label, absence of placebo, the inclusion of suspected (10%) and laboratory-confirmed (90%) cases (post-hoc analysis in confirmed cases yielded similar results), and absent multiple testing adjustment, block randomization, and pre-specification rules. Arrhythmias were not different (44.7% vs. 43%); one spontaneously resolved torsades des pointes SAE occurred with HC.

The FDA revoked its March 2020 EUA for HC/CQ for hospitalized patients with COVID-19 (outside trials) on June 15, 2020, based on results of the *Recovery Trial* and other emerging data [7].

An NIH press release on June 20, 2020, announced the final permanent cessation of enrollment in its *ORCHID* trial for futility after a fourth interim analysis showed no mortality benefit (albeit minimal safety issues). This (third) RCT provided the first robust, blinded, and placebo-controlled (not open-labeled) data for HC in hospitalized patients. Eventually published in JAMA on November 9, 2020, 479 patients were enrolled from 34 U.S. sites with a median interval from symptoms onset of 5 days (relatively early) [28]. Corticosteroids and azithromycin use was similar in the two treatment groups. The primary outcome measurement, the WHO 14-day ordinal score was similar in HC vs. placebo patients (median [IQR] score, 6 [4-7] vs 6 [4-7]; aOR, 1.02 [95% CI, 0.73 to 1.42]). No differences in secondary outcomes (including mechanical ventilation) – or mortality were found (10.4% vs. 10.6%, respectively) (absolute difference, −0.2% [95% CI, −5.7% to 5.3%]; aOR, 1.07 [95% CI, 0.54 to 2.09]). QTc prolongation was more common with HC (5.9% vs 3.3%) but SAE rates were similar (5.8% vs. 4.6%).

A WHO press release on July 4, 2020, announced final permanent discontinuation of enrollment in its *Solidarity* Trial HC arm after interim analysis similarly showed no mortality benefit but concerning safety signals (fourth RCT).^(38)^ Results were eventually published in NEJM on December 2, 2020 [29]. The primary outcome, intention-to-treat in-hospital mortality, in this large open-labeled RCT at 405 hospitals in 30 countries, occurred in 104 of 947 (11.0%) HC vs. 84 of 906 (9.3%) control patients (rate ratio, 1.19; 95% CI, 0.89-1.59, p=0.23); neither need for mechanical ventilation nor hospital length of stay were significantly reduced by HC. The trial’s open-label design without placebo is an obvious limitation but is unlikely to have biased mortality results.

The fifth spring/summer 2020 RCT, the *Coalition Covid-19 Brazil I* study [30] was multicenter, randomized, open-label, and controlled, comparing HC with or without azithromycin and standard care in 504 laboratory confirmed mild-moderate (requiring </= 4 L oxygen) hospitalized COVID-19 patients. The median interval from symptoms onset was 7 days. No differences were found for the primary outcome, clinical assessment at 15 days (seven-level ordinal score), comparing the treatment groups vs. the standard care group in a modified ITT analysis (confirmed cases only) (HC vs. standard care, OR 1.21; 95% CI, 0.69-2.11; p=1.00; HC with azithromycin vs. standard care, OR 0.99, 95% CI 0.57-1.73; p=1.00). QTc prolongation and LFTs elevation were more frequent with HC. Trial limitations consisted of open-design without placebo, smaller size, and restriction to mild-moderate severity.

Updated NIH and Infectious Diseases Society of America (IDSA) COVID-19 guidelines in June 2020 recommended HC/CQ use in hospitalized COVID-19 patients only in clinical trials; NIH (27 August 2020) and IDSA (20 August 2020) guidelines were then extended to an emphatic recommendation against HC/CQ use in hospitalized COVID-19 patients [67-68].

At least 12 additional RCTs were published evaluating HC/CQ in hospitalized COVID-19 patients published later on in the pandemic (winter of 2020, and in 2021). All showed similar absence of convincing evidence of benefit, and some worse primary outcome measurements (including clinical ordinal scales, mortality, composites, and viral shedding); some showed concerning safety signals including higher QTc prolongation, renal injury, and AE/SAE rates [31-42]. Outcomes from these RCTs are briefly summarized below.

A small RCT in 53 patients showed no difference in viral clearance between HC and HC/SOC treated patients [31]. An Egyptian RCT in 194 patients showed no difference in need for mechanical ventilation or mortality between HC and HC/SOC patient [32]. In New York University’s TEACH double-blinded RCT in 128 patients comparing HC and placebo, there was no difference in the study’s primary outcome, severe disease progression composite end point; viral clearance and AE rates were similar in the two treatment groups [33]. In Intermountain’s HAHPS RCT comparing 85 patients treated with HC vs. azithromycin with a Bayesian analysis, no convincing difference was found for the primary outcome, the 14-day ordinal score [34]; AE rates, QTc prolongation were similar but the AKI rate was numerically higher with HC. In a combined report of a Taiwanese small open-labeled RCT (n=34) and small retrospective study (n=37), 14-day viral clearance was similar with or without HC [35]. In a Brazilian open-labeled RCT in 105 patients, addition of HC or CQ to SOC resulted in significant worsening of the primary outcome, a 14-day 9-point clinical ordinal score, as well as need for mechanical ventilation and severe AKI (but not arrhythmias) [36]. In another Brazilian RCT in 168 patients randomized to receive HC, CQ, or ivermectin, there were no significant differences in the primary endpoints, need for oxygen or mechanical ventilation, ICU admission, or mortality [37]. In the REMAP-CAP trial in patients were treated with lopinavir-ritonavir (n=255), HC (n=50), combination therapy (n=27) or control (n=362), a Bayesian analysis of its primary endpoint of an ordinal scale of organ support-free days (as well as mortality) showed significantly worse outcome with all 3 treatments vs. control [38]. In HYCOVID, a double-blinded RCT in 247 patients with milder disease in France and Monaco comparing HC and placebo, neither the primary outcome of a 14-day composite of death and need for mechanical ventilation, nor viral clearance, were different [39]. In a Mexican double-blinded RCT in 214 patients comparing HC and placebo, neither the primary outcome, 30-day mortality, nor any secondary outcomes differed [40]. in NOR-Solidarity, a Norwegian add-on study to WHO’s *SOLIDARITY* trial, viral clearance, respiratory failure severity, and inflammatory variables were compared (and in-hospital mortality) in 185 patients receiving remdesivir, HC, or SOC. There were no group differences for any of these variables [41]. In a Danish double-blinded, placebo-controlled RCT in 117 patients, the primary outcome, days alive and discharged from hospital by 14 days, did not differ between HC/azithromycin vs. placebo/placebo [42]. See Table 1.

**Table 1.** RCTs evaluating HC/CQ in hospitalized patients with COVID-19.

| Reference | N | Design | Comparators | Primary outcome | Primary results | Other results |
| --- | --- | --- | --- | --- | --- | --- |
| Tang W [26] | 150 | Open-label | HC+SOC vs. SOC | Viral clearance | No significant difference | Numerically more AEs with HC but no arrhythmias or QTc prolongation. |
| Horby PW [27]<br>*RECOVERY* | 4,716 | Open-label, platform | HC+SOC vs. SOC | 28-day mortality | No significant difference | Higher composite endpoint of mechanical ventilation requirement & death with HC. Arrhythmias were similar. |
| Self W [28]<br>*ORCHID* | 479 | Double-blinded, placebo-controlled | HC+SOC vs. Placebo+SOC | WHO 14-day ordinal score | No significant difference | Mortality and mechanical ventilation need were similar. QTc prolongation was numerically higher with HC but SAEs were similar. |
| Pan H [29]<br>*SOLIDARITY* | 1,853 | Open-label, platform | HC+SOC vs. SOC | In-hospital mortality | No significant difference | Mechanical ventilation need & LOS were similar. |
| Cavalcanti AB [30]<br>*Coalition Covid-19 Brazil I* | 504 | Open-label, mild-moderate severity | HC+SOC vs. SOC & HC+AZ+SOC vs. SOC | 15-day ordinal score | No significant differences | QTc prolongation & elevated LFTs were numerically higher with HC. |
| Lyngbakken MN [31] | 53 | Open-label | HC+SOC vs. SOC | Viral clearance | No significant difference | Mortality, ordinal score, & LOS were similar. |
| Abd-Elsalam S [32] | 194 | Open-label | HC+SOC vs. SOC | Need for mechanical ventilation or death | No significant differences |  |
| Ulrich RJ [33]<br>*TEACH* | 128 | Double-blinded, placebo-controlled | HC+SOC vs. Placebo+SOC | Composite of severe disease progression | No significant difference | Viral clearance & AEs were similar. |
| Brown SM [34]<br>*HAHPS* | 85 | Open-label, AZ control, Bayesian analysis | HC vs. AZ | 14-day ordinal score | No significant difference | AEs & QTc prolongation were similar but AKIs were numerically higher with HC. |
| Chen CP [35] | 34 | Open-label | HC+SOC vs. SOC | Viral clearance | No significant difference |  |
| Rea-Neto A [36] | 105 | Open-label | HC/CQ+SOC vs. SOC | 14-day ordinal score | Significantly worse with HC | Mechanical ventilation need and severe AKIs were significantly higher with HC. Arrhythmias were similar. |
| Galan LEB [37] | 168 | Double-blinded, ivermectin-controlled | HC vs. CQ. vs. ivermectin | Need for O2 or mechanical ventilation, ICU admission, or mortality | No significant differences | SAEs were similar. |
| Arabi YM [38]<br>*REMAP-CAP* | 694<br>L-R(N=255)<br>HC (N=50)<br>L-R & HC (N=27)<br>SOC (N=362) | Open-label, platform, Bayesian analysis | Lopinavir-ritonavir+SOC vs. HC+SOC vs. lopinavir-ritonavir & HC+SOC vs. SOC | Ordinal scale of organ support-free days | Significantly worse with all 3 treatments | Significantly worse mortality with all 3 treatments. |
| Dubee V [39]<br>*HYCOVID* | 247 | Double-blinded, placebo-controlled | HC+SOC vs. Placebo+SOC | 14-day composite of death & need for mechanical ventilation | No significant difference | Viral clearance, ordinal scores, & AEs & SAEs were similar. |
| Hernandez-Cardenas C [40] | 214 | Double-blinded, placebo-controlled | HC+SOC vs. Placebo+SOC | 30- day mortality | No significant difference | Need for mechanical ventilation, LOS, & SAEs were similar. |
| Barratt-Due A [41]<br>*NOR-Solidarity* | 185 | Open-label, platform, add-on to *SOLIDARITY* | HC+SOC vs. remdesivir+SOC vs. SOC | Viral clearance | No significant difference | Respiratory failure severity, inflammatory variables, & in-hospital mortality were similar. |
| Sivapalan P [42] | 117 | Double-blinded, placebo-controlled | HC/AZ+SOC vs. Placebo/Placebo+SOC | Days alive & discharged from hospital by 14 days | No significant difference | Diarrhea was numerically higher with HC/AZ but QTc prolongation and SAEs were numerically higher with Placebo. |

A prospective controlled but unrandomized trial in 66 patients in Brazil also failed to show a difference in viral clearance [69].

An interesting and telling study from Israel found, for therapeutics in COVID-19, low concordance between published observational studies (often ‘positive’) and RCTs (usually ‘negative), akin to findings in this HC/CQ review [70].

Key limitations of these 17 RCTs include moderate risk of bias in 13 (majority) and optimal double-blinded placebo-controlled design in only 5 (minority).

### Meta-analyses in Hospitalized Patients

In the overwhelming majority of 50 published aggregate date meta-analyses (AD-MAs) and in our IPD-MA [45] assessing HC/CQ in hospitalized COVID-19 patients, no convincing efficacy was found, and in many adverse safety signals were noted. Ten of these reported limited patient level subgroup analyses with a pattern of subgroup results paralleling overall results. Three AD-MA examples follow:

A large *Open Society Foundation* AD-MA from 14 published and 14 unpublished HC (26 trials) and CQ (4 trials) studies was published; 67% of the sample size of 10,319 patients was derived from the *RECOVERY* and *SOLIDARITY* trials [43]. Mortality was found to be higher with HC (OR 1.11 [95% CI: 1.02-1.20; 26 trials; 10,012 patients) and equivocal for CQ (OR 1.77 [95% CI: 0.15-21.13, 4 trials; 307 patients). Patient level subgroup analysis was restricted to disease severity.

A Cochrane AD-MA from 12 RCTs with 8,569 COVID-19 patients [44]. No differences were found for HC/CQ vs. control treatment for mortality (RR 1.09, 95% CI 0.99-1.19), mechanical ventilation (RR 1.11, 95% CI 0.91-1.37), or conversion to negative nasopharyngeal swabs (RR 1.00, 95% CI 0.91-1.10); AEs were more frequent with treatment (RR 2.90, 95% CI 1.49-5.6) but not SAEs (RR 0.82, 95% CI 0.37-1.79). Subgroup analyses were planned but no completed due to inability to secure necessary data.

Results of an IPD-MA of 8 U.S. RCTs evaluating HC/CQ in hospitalized COVID-19 patients also showing no credible efficacy in hospitalized COVID-19 patients in the overall population (OR 0.95; 95% credible interval 0.77 to 1.22) and in a comprehensive analysis of multiple patient level subgroups (NCOSS (a disease severity surrogate), age, gender, number of comorbidities, BMI, and estimated baseline risk) was recently submitted for publication [45]. Overall AE, SAE, and elevated LFTs AE rates were numerically higher with HC/CQ but not QTc prolongation or arrhythmias.

### Clinical Studies in Other Indications

#### Outpatients

A retrospective-observational trial from New Jersey comparing outcome in 1274 HC treated outpatients with COVID-19 and 1067 patient propensity-matched cohort, hospitalization occurred in 21.6% vs. 31.4%, respectively (OR 0.53; 95% CI, 0.29-0.95) [71].

Akin to results in inpatients (above), RCTs failed to show HC/CQ efficacy in outpatients:

A double-blinded and placebo-controlled RCT was published evaluating HC in 423 COVID-19 outpatients (81% laboratory-confirmed or exposed to a laboratory-confirmed individual) with mild disease [72]. Change in a symptom severity score over 14 days, the study’s primary outcome measurement, was not statistically different, nor were hospitalization rates. Mild adverse events were more common with HC.

Another RCT in COVID-19 outpatients, Q-PROTECT, randomized patients with mild-moderate symptoms to placebo or HC with or without azithromycin. In the 456 patients enrolled, viral cure (PCR negativity at day 6), the primary outcome measurement, was similar in all three groups (12.2%, 10.5%, and 12.8%, respectively; p=0.821) [73].

In another RCT in COVID-19 outpatients (all were laboratory-confirmed) in Alberta, HC and placebo were compared. Treatment occurred at a mean of 12 days after symptoms onset. The primary outcome, a composite of 30-day hospitalization/mechanical ventilation/death occurred in 4 of 111 randomized HC patients (4 hospitalizations) vs. 0 of 37 placebo patients. Symptoms duration was not decreased either. The study was terminated prematurely due to slow recruitment [74].

A RCT from Brazil compared outcome in 685 COVID-19 outpatients randomized to receive HC, lopinavir/ritonavir, or placebo. The primary outcome, hospitalizations, occurred in 3.7% vs. 5.7% vs. 4.8%, respectively (HR 0.76, 95% CI, 0.30-1.88]. There were no secondary outcomes group differences either (mortality, viral shedding) [75].

A small placebo-controlled RCT in 84 outpatients found no difference in 9-day viral clearance between HC and placebo [76].

In summary, HC/CQ has not been found to have significant efficacy in outpatients with COVID-19 yet increased AEs in some studies, however the published database is more limited than for hospitalized patients.

#### Post exposure prophylaxis

A post-exposure prophylaxis study was reported in which 821 subjects with moderate- or high-risk household or occupation exposure were randomized to receive HC or placebo within 4 days of exposure [77-78]. The primary outcome, “*incidence of new illness compatible with Covid-19*” (fewer than 3% were laboratory confirmed) was not different (11.8% vs. 14.3%, p=0.35). None-serious AEs were more common with HC.

Another post-exposure prophylaxis study in 2,314 with an open-label cluster-randomization design found the primary outcome of PCR-confirmed symptomatic COVID-19 infection to be similar in HC vs. usual care patients (5.7% and 6.2%, respectively; risk ratio, 0.86 [95% confidence interval, 0.52 to 1.42]); non-serious adverse events were much more common with HC (56.1% vs. 5.9%) but no ‘related’ SAEs occurred [79].

In another post-exposure prophylaxis study, household exposures within 96 hours were randomized to receive HC (n=407) or vitamin C placebo (n=334). Among 689 participants who were PCR swab at baseline, conversion to positive PCR (the primary outcome) occurred 53 vs. 45 subjects (adjusted HR, 1.10, 95% CI, 0.73-1.66; p>0.2). The AE rate was significantly higher with HC (16.2 vs. 10.9%, p=0.026) [80].

In summary, HC/CQ has not been found to have significant efficacy for pre-exposure prophylaxis against COVID-19 yet increased AEs, however the published database is more limited than for hospitalized patients.

#### Pre-exposure prophylaxis

Four pre-exposure prophylaxis studies (two double-blind placebo-controlled RCTs in healthcare workers and two large observational retrospective population-based studies in rheumatoid arthritis and lupus patients) showed no differences in COVID-19 infection rates [81-83] or mortality [84]. A descriptive safety analysis from three of these outpatient RCTs (1 non-hospitalized mild-moderate disease RCT, 1 post-exposure prophylaxis RCT, and 1 pre-exposure prophylaxis RCT) in 2,795 subjects found increased AE rates with HC (36-40%) vs. placebo (19%) but rare SAEs; GI upset was the most common AE. Co-administration of other QT prolonging medications was an exclusion in these RCTs [85].

An open-label cluster study in migrants in Singapore showed higher viral clearance with HC vs. vitamin C control [86]. An Indian open-labeled, controlled study in 317 exposed or presumed exposed subjects showed significantly decreased infection rates with HC post-exposure prophylaxis vs. SOC [87].

A potential critique of some of the post-exposure prophylaxis studies has been that HC/CQ was sometimes administered late after symptoms onset leading to decreased efficacy – as occurs with delayed neuraminidase inhibitor treatment for influenza and in some experimental SARS-CoV-2 mouse models [88]. Higher dosing than necessary based on predicted pharmacokinetics, leading to more adverse events, has also been a critique.

An unpublished medRxiv aggregate date meta-analysis including five pre- and post-exposure prophylaxis RCTs reported possible benefit for HC [89].

In summary, HC/CQ has not been found to have significant efficacy for pre-exposure prophylaxis against COVID-19 in most but not all studies and increased AEs in some, however the published database is much more limited than for hospitalized patients.

### Study limitations

The key limitation of this review is that systematic adherence to PRISMA checklist [90] components was high for HC/CQ studies in hospitalized patients (the focus of this review) but lower for the other studies (e.g., no risk of bias assessment in the latter).

### Conclusions

This systematic review of preclinical *in vitro* and animal studies, retrospective-observational trials, RCTs, and meta-analyses strongly suggests that HC/CQ are ineffective in hospitalized patients with COVID-19, both in overall populations and in subpopulations, and should not be administered outside of RCTs with robust informed consent about unlikely benefit and probable harm. We believe that the preclinical and clinical database was never sufficient to support empiric use.

The published clinical trials database for HC/CQ in outpatients and post-exposure and pre-exposure prophylaxis also shows lack of convincing efficacy despite increased adverse events, but it is more limited than for hospitalized patients, particularly for pre-exposure prophylaxis. Our review was less robust for these indications.

Empiricism, particularly when based mainly on retrospective-observational studies rather than RCTs, is fraught with danger for patients. Cognizance of the story of HC/CQ’s failure during the COVID-19 pandemic should lead to refraining on the part of the medical community from repeating the same errors for other experimental therapeutics. Dr. Kalil’s wise admonition in his May 2020 *JAMA* viewpoint bears repeating: “*The administration of any unproven drug as a ‘last resort’ wrongly assumes that benefit will be more likely than harm*” [91].

## Data Availability

All data produced in the present work are contained in the manuscript.

## Acknowledgments

We thank Dr. Kunjal Luhadia, MD, for her helpful manuscript edits.

## Funding Source

None.

## Notes

### Competing Interest Statement

The authors have declared no competing interest.

### Funding Statement

This study did not receive any funding.

