## Supplementary material for "Hydroxychloroquine/Chloroquine in COVID-19 With Focus on Hospitalized Patients – A Systematic Review": Table S1

**Table S1. RCTs evaluating HC/CQ in hospitalized patients with COVID-19 – risk of bias assessment**

| Reference | Random sequence generation | Allocation concealment | Blinding of participants and personnel | Blinding of outcome assessments | Incomplete outcome data | Selective reporting | Other bias | Overall bias assessment |
|---|---|---|---|---|---|---|---|---|
| Tang W [26] | Low | Low | High | Moderate | Moderate | Low | Low | Moderate |
| Horby PW [27] *RECOVERY* | Low | Low | High | Moderate | Moderate | Low | Low | Moderate |
| Self W [28] *ORCHID* | Low | Low | Low | Low | Moderate | Low | Low | Low |
| Pan H [29] *SOLIDARITY* | Low | Low | High | Moderate | Moderate | Low | Low | Moderate |
| Cavalcanti AB [30] *Coalition Covid-19 Brazil I* | Low | Low | High | Moderate | Moderate | Low | Low | Moderate |
| Lyngbakken MN [31] | Low | Low | High | Moderate | Moderate | Low | Low | Moderate |
| Abd-Elsalam S [32] | Low | Low | High | Moderate | Moderate | Low | Low | Moderate |
| Ulrich RJ [33] *TEACH* | Low | Low | Low | Low | Moderate | Low | Low | Moderate |
| Brown SM [34] *HAHPS* | Low | Low | High | Moderate | Moderate | Low | Low | Moderate |
| Chen CP [35] | Low | Low | High | Moderate | Moderate | Low | Low | Moderate |
| Rea-Neto A [36] | Low | Low | High | Moderate | Low | Low | Low | Moderate |
| Galan LEB [37] | Low | Low | Low | Low | Moderate | Low | Low | Low |
| Arabi YM [38] *REMAP-CAP* | Low | Low | High | High | Low | Low | Low | Moderate |
| Dubee V [39] *HYCOVID* | Low | Low | Low | Low | Low | Low | Low | Moderate |
| Hernandez-Cardenas C [40] | Low | Low | Low | Low | Moderate | Low | Low | Low |
| Barratt-Due A [41] *NOR-Solidarity* | Low | Low | High | Moderate | Moderate | Low | Low | Moderate |
| Sivapalan P [42] | Low | Low | Low | Low | Moderate | Low | Low | Low |

The Cochrane Collaboration tool was used to assess risk of bias [92], one assessor was used, lack of blinding led to 'high risk' assessments for the blinding domains; overall bias assessments were usually moderate anyhow because outcome measurements were 'hard' and less likely to be affected by lack of blinding. Absent notation of how missingness was handled resulted in a moderate risk domain assessment for 'incomplete outcome data'.
